# Modeling COVID 19 in the Basque Country: from introduction to control measure response

**DOI:** 10.1101/2020.05.10.20086504

**Authors:** Maíra Aguiar, Eduardo Millán Ortuondo, Joseba Bidaurrazaga Van-Dierdonck, Javier Mar, Nico Stollenwerk

## Abstract

In March 2020, a multidisciplinary task force (so-called Basque Modelling Task Force, BMTF) was created to assist the Basque Health managers and the Basque Government during the COVID-19 responses. BMTF is a modeling team, working on different approaches, including stochastic processes, statistical methods and artificial intelligence. In this paper we describe and present the results obtained by a new stochastic SHARUCD model framework which was able to describe the disease incidence data provided by the Basque Health Services. Our models differentiate mild and asymptomatic from severe infections prone to be hospitalized and were able to predict the course of the epidemic, from introduction to control measure response, providing important projections on the national health system necessities during the increased population demand on hospital admissions. Short and longer-term predictions were tested with good results adjusted to the current epidemiological data, showing that the partial lockdown measures were effective and enough to slow down disease transmission in the Basque Country. The growth rate *λ* Is calculated from the model and from the data and the implications for the reproduction ratio *r* are shown. At the moment, the reproduction ratio *r* is estimated to be below the threshold behavior of *r* = 1, but still close to 1, meaning that although the number of new cases are decelerating, a careful monitoring of the development of the outbreak is required. This framework is now being used to monitor disease transmission while the country lock-down is gradually lifted, with insights to specific programs for a general policy of “social distancing” and home quarantining. These are the first publicly available modeling results for the Basque Country and the efforts will be continued taking into consideration the updated data and new information that are generated over time.

## 1 Introduction

In December 2019, a new respiratory syndrome (COVID-19) caused by a new coronavirus (SARS-CoV-2), was identified in China [1] and spread rapidly around the globe. With human-to-human transmission confirmed in 3 countries outside China, COVID-19 was declared a Public Health Emergency of International Concern by the World Health Organization (WHO) on 30 January 2020. By February 25th, 2020, China was the epicenter of the outbreak and, in 2 weeks, on March 11th, 2020, COVID-19 was characterized as a pandemic, with Europe reporting more cases and deaths than the rest of the world combined, apart from China [2]. Up to April 25th, 2020, more than 2.8 million cases were confirmed with about 200 thousand deaths, with a global case fatality ratio (CFR) of approximate 7% [3].

Italy, the first hardest hit country in Europe, had local transmission confirmed in all regions in the beginning of March, 2020. Eleven municipalities in northern Italy were identified as the centers of the two main Italian clusters and, on March 8th, 2020, the Prime Minister Giuseppe Conte has placed in quarantine all of Lombardy and 14 other northern provinces. The national lockdown decree was signed on March 9, 2020 [4], prohibiting all forms of gathering of people in public places and suspending sports events and competitions of all types. By that time, Italy, was considered the new epicenter of the outbreak, reporting more than 9 thousand confirmed cases with more than 450 deaths. On March 21, 2020, further restrictions within the nationwide lockdown were imposed with all non-essential production, industries and businesses halted, as the number of new cases and deaths were still rising. In respect to the total number of confirmed cases, Spain was 8 days behind Italy, with cases reported in all 50 provinces of the country on March 8, 2020. The decree of a national lockdown was signed on March 14, 2020 [5], with all non-essential workers staying at home from March 27th, 2020 [6, 7] on.

In the Basque Country, an autonomous community in northern Spain with 2.2 milion inhabitants, the first cases of COVID-19 were notified on March 4th, 2020. A public health emergency was declared before any other region in Spain [8]. All schools in the Basque Country were closed by March 12, 2020, and, ruled by the same Spanish decrees [5, 6, 7], lockdown measures were implemented accordingly and in time. An extension of the state of alarme was published on April 10th, 2020 [9], and although teleworking is still prioritized, some restrictions started to be lifted, with workers in some non-essential sectors allowed to return to work using face masks on April 13, 2020 and children under the age of 14 allowed to go outside for a walk, within a one-kilometer radius of their home, on April 26, 2020 [10]. The national plan for lifting the restrictions imposed during the state of alarm called “Plan for the Transition towards a new normality” was announced on April 28, 2020 [11] and will take place over 4 phases with a “gradual, flexible and adaptive” de-escalation to “a new normality”, depending on the on-going progress of COVID-19 epidemic’s control across the different regions of Spain. Started on May 4, 2020, with its “Phase Zero”, the proposed plan will last eight weeks, until the end of June.

As the COVID-19 pandemic is unfolding, research on mathematical modeling becomes more important than ever to understand disease spreading dynamics and the impact of intervention measures. By incorporating the new information generated by virology, field epidemiology and social behaviour, for example, mathematical models are often used to guide public health authorities with projections for the national health systems needs during an outbreak. Those mathematical models also provide insights about the disease spreading over time, assessing the impact of human interventions for disease control, and Governments in some countries have already taking important decisions based on these results [12, 13, 14]. Worldwide country lockdowns are unprecedented draconian measures recently taken and, although needed to decelerate disease transmission, have caused a huge economical crises around the globe. As some countries on the northern hemisphere start to announce that they were able to control the spreading of the disease and have now “reached the peak” of the epidemic, Governments start to consider to relax the imposed restrictions, and once again, mathematical models become essential guiding tools to evaluate the impact of the ongoing partial lockdowns lifting on disease transmission intensities, for different epidemiological scenarios, combined with many other public health measures that must take place for continuing COVID-19 prevention and mitigation such as testing, contact tracing and isolation of infected individuals.

The COVID-19 pandemic has resulted in an avalanche of epidemiological modeling papers, most of them using simple models such as the SIR (Susceptible-Infected-Recovered) or SEIR (Susceptible-Exposed-Infected-Recovered, another framework used withing the BMTF) [15] in mechanistic or probabilistic frameworks to understand and predict the spread of the disease in a population. With valuable results, modeling the dynamics of COVID-19 is very challenging, as we know very little about the disease. More complex models would be able to give more accurate projections about specific variables such as number of hospitalizations, intensive care units admissions (ICUs) and deaths, for example, over the course of the epidemics. However, to build useful models, good quality empirical data and its understanding, as well as a close collaboration among mathematical modelers, field and laboratory researchers as well as public health stakeholders are essential [16, 17, 18, 19].

In March 2020, a multidisciplinary task force (so-called Basque Modelling Task Force, BMTF) was created to assist the Basque Health managers and the Basque Government during the COVID-19 responses. BMTF is a modeling team, working on different approaches, including stochastic processes, statistical methods and artificial intelligence. Members were collaborating taking into consideration all information provided by the public health frontline and using different available datasets in respect to the COVID-19 outbreak in the Basque Country. The objectives were, besides projections on the national health system needs during the increased population demand on hospital admissions, the description of the epidemic in terms of disease spreading and control, as well as monitoring the disease transmission when the country lockdown was gradually lifted. All modeling approaches were complementary and were able to provide coherent results, assuring that the decisions made using the modeling results were sound and, in fact, adjusted to the current epidemiological data. In addition, modeling results provided useful predictive information to validate outbreak control decisions and finally, to assist authorities in the Basque Country. In this paper we describe and present the results obtained by one of the modeling approaches developed within the BMTF, specifically using extended versions of the basic epidemiological SIR-type models, able to describe the dynamics observed for tested positive cases, hospitalizations, intensive care units (ICUs) admissions, deceased and finally the recovered. Keeping the biological parameters for COVID-19 in the range of the recent research findings [20, 21, 22, 23, 27, 28], but adjusting to the phenomenological data description, the models were able to explain well the exponential phase of the epidemic and fixed to evaluate the effect of the imposed control measures. With good predictability so far, consistent with the updated data, we continue this work while the imposed restrictions are relaxed and closely monitored.

We use stochastic SHARUCD-type models (susceptible (S), severe cases prone to hospitalization (H), mild, sub-clinical or asymptomatic (A), recovered (R), patients admitted to the intensive care units (U) and the recorded cumulative positive cases (C) which includes all new positive cases for each class of H, A, U, R, and deceased (D)) – an extension of the well known simple SIR model that is frequently used to model different disease outbreaks [24, 25, 26]. The deterministic approach is obtained via the mean field approximation and both frameworks are used to evaluate the model performance and accuracy. The model is calibrated using the empirical data for the Basque Country community and the biological parameters are estimated and fixed as the model is able to describe the disease incidence data. The growth rate (*λ*) is calculated from the model and from the data and results for the reproduction ratio (*r*) are shown. As the epidemic has entered into its linear phase, with the new number of cases increasing less and stabilizing, we now monitor the effect of the control measures on disease transmission and give a longer-term prediction of the development of the epidemic, for the present epidemiological scenario, towards its control. Preparation for a possible second wave of transmission are also discussed, as the influence of seasonality and the extend of acquired immunity and its duration against SARS-CoV-2 are not clear nor well measurable yet. This will be specially important when the lockdown is completed lifted.

## 2 Methods and Results

### 2.1 Epidemiological Data and data inspection

Epidemiological data used in this study are provided by the Basque Health Department and the Basque Health Service (Osakidetza), continually collected with specific inclusion and exclusion criteria, and for the present analysis, the last update was on May 4, 2020. This is a dynamical work and new results are presented throughout the manuscript as new data are collected to calibrate the models. We use the following incidence and cumulative data for RT-PCR (real time polymerase chain reaction) tested positive patients (yellow), recorded as hospital admissions (red), intensive care units admissions (purple), recovered (green) and deceased (black). The remainder are assumed to be individuals with milder infections. Data sets for hospitalizations, ICU admissions, discharges and deceases were obtain from the Osakidetza’s Business Intelligence (OBI) platform, enabling us to exploit and analyze the empirical data from the main information systems of Basque Health Service, including the structured data from the electronic health records of the Basque Health Service.

At the beginning of the outbreak, RT-PCR tests were only performed to those patients with severe symptoms admitted to a hospital. From March 22 on, testing capacities has increased, with antibody tests also available and used mainly as screening tool in nursing homes, with less severe symptomatic cases started to be tested. Data from different sources are linked by a pseudonymous identification of the patient.

#### 2.1.1 Definition of each variable and data flow path withing the BMTF

Positives cases are patients tested positive for the first time with a RT-PCR test. Hospitalizations and ICUs refer to tested positive patients admitted to a hospital and intensive care unit admissions. A unique episode (hospitalization) is considered in case of patient transfer between hospitals in the Basque Country. To avoid including hospitalizations not related to COVID-19 infection, hospitalizations with a discharge data before the notification of the positive test are excluded and only hospitalizations that at a certain moment were in charge of services a priori responsible of attending covid disease are included: (internal medicine, pediatrics, ICU or reanimation services, respiratory services and infection disease services). Recovered variable are referring notified hospital and ICU discharges, excluding death. This data set does not include recovered individuals that were tested positive but not admitted to a hospital and like that represents only part of the expected recoveries in the population. Deceased include both hospital and outside hospital deaths. Data from different sources are linked by a pseudonymous identification of the patient.

To validate those variable definitions with the available data, the following path was used: i) data are extracted from OBI and transformed following the above described definitions. ii) data sets are shared with collaborators from different health organizations to compare and check the data collection and to eventually revise variable definitions, adjusting if necessary. iii) data are used to validate mathematical models developed to describe COVID-19 dynamics in the Basque Country and results are discussed during regular BMTF meetings with regular reports delivered to the Basque Health managers and the Basque Government. iv) feedback on the reports are sent to the researchers working with data with adjustments on models parameters and data collection when necessary.

As first data inspection, the dynamics of the cumulative cases together with the effective starting dates for different control measures imposed are shown in Fig. 1.

**Figure 1:**
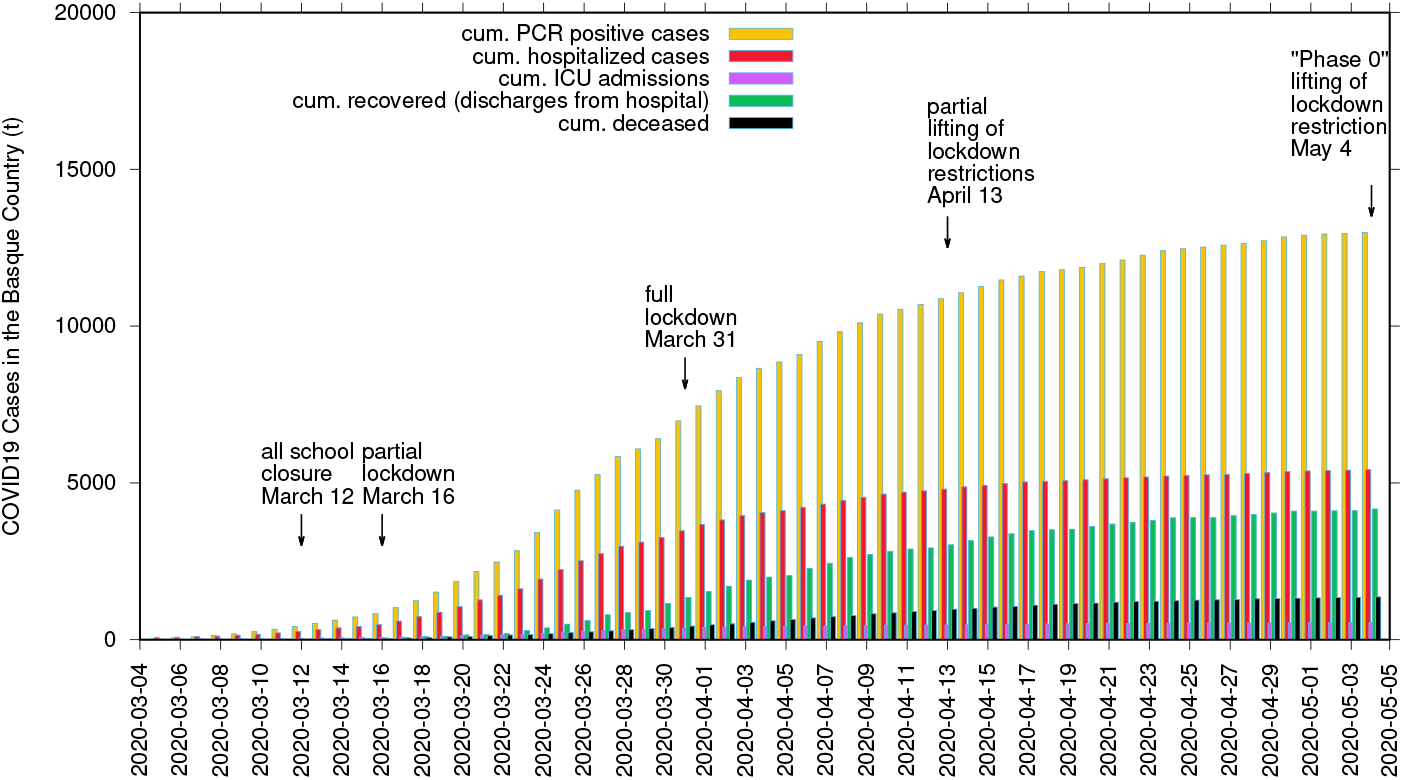
Cumulative COVID-19 cases of tested positive (*I*_*cum*_), hospitalized cases (*C*_*H*_), ICU admission (*C*_*U*_), recovered (*C*_*R*_) and deceased cases (*D*). The imposed control measures and its gradual lifting process are marked with arrows with the effective dates of implementation.

### 2.2 Modeling framework

We use SHARUCD-type models, an extension of the well known simple *SIR* (susceptible-infected-recovered) model, with infected class *I* partitioned into severe infections prone to hospitalization (*H*) and mild, sub-clinical or asymptomatic infections (*A*). From a typical SIR model with constant population size *N* = *S* + *I* + *R*, infection rate *β* and recovery rate *γ* (and an eventual waining immunity rate *α*, which in the case of COVID-19 is not yet relevant as we assume, preliminarily, that infection leads to immunity during the time horizon considered up to now) given by

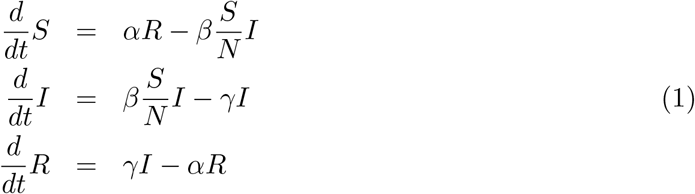

we develop a basic SHAR-model including now a severity ratio *η* for susceptible individuals (*S*) developing severe disease and possibly being hospitalized *H* or (1 − *η*) for milder disease, including sub-clinical and eventually asymptomatic *A* infections, where mild infected *A* have different infectivity from severe hospitalized disease *H*, parametrized by a ratio *ϕ* to be smaller or larger than *ϕ* = 1 comparing to baseline infectivity rate *β* of the “hospitalized” *H* class and altered infectivity rate *ϕ · β* for mild/“asymptomatic” *A* class. Hence we obtain a SHAR-type model, where parameters for disease induced death (*µ*) and intensive care units admission (*ν*) are indicated, being relevant for severe disease prone to hospitalization but not for mild/asymptomatic cases, reads

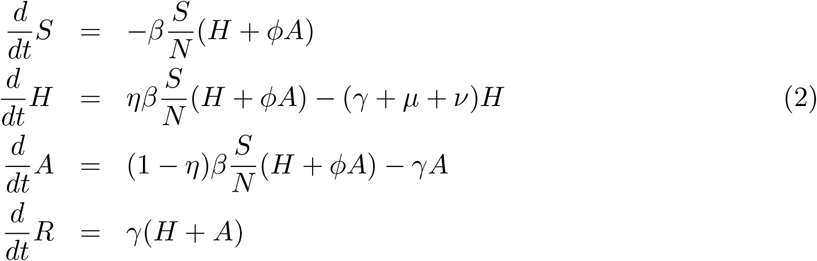

that needs to be further refined to describe COVID-19 dynamics. Such model sketch needs to be further refined adjusting the modelling framework to the available empirical data. For investigation of *ϕ* to be around one or even larger due to higher mobility of mild infected or asymptomatic see e.g. [43].

For severe infections prone to hospitalization, we assume the following dynamics: severe hospitalized individuals *H* could either recover, with a recovery rate *γ*, be admitted to the ICU facilities *U*, with a rate *ν*, or eventually decease into class *D* before being admitted to the ICU facilities, with a disease induced death rate *µ*. The ICU admitted patients could recover or die. For completeness of the system and to be able to describe the initial introductory phase of the epidemic, an import term *ϱ* should be also included into the force of infection. For the present study, we assume *ϱ* to be much smaller than the other additive terms of the force of infection, given the strong observational insecurities on the data collected at the beginning of the outbreak, when *ϱ* would matter most.

As we investigate cumulative data on the infection classes and not prevalence, we also include classes *C* to count cumulatively the new cases for “hospitalized” *C*_*H*_, “asymptomatic” *C*_*A*_, recovered *C*_*R*_ and ICU patients *C*_*U*_. In this way we can easily include a ratio *ξ* of under- notification of mild/asymptomatic cases. The deceased cases are automatically collecting cumulative cases, since there is no exit transition form the death class *D*. While *H* and *U* can describe the empirical data reported for each class, the recovered *R*, which are also cumulative, count all biologically recovered, including the undetected mild/asymptomatic cases. Therefore, the *C*_*R*_ class is needed to describe the present data which count only the notified asymptomatic in terms of *ξA*. We have finally the SHARUCD-type models, where individual transitions are still subject to refinement upon information obtained about COVID-19 biological mechanisms and on additional information directly obtained from the data. For a basic first SHARUCD model we keep a balance between biologically necessary and relevant model classes, and transitions, and a possibly relatively low number of free parameters, able to be estimated with the presently available data, avoiding over-parametrization as much as possible.

We consider primarily SHARUCD model versions as stochastic processes in order to compare with the available data which are often noisy and to include population fluctuations, since at times we have relatively low numbers of infected in the various classes. The stochastic version can be formulated through the master equation [34, 35, 36] in application to epidemiology [29, 40] in a generic form using densities of all variables *x*_1_ := *S/N, x*_2_ := *H/N, x*_3_ := *A/N, x*_4_ := *R/N, x*_5_ := *U/N, x*_6_ := *C*_*H*_*/N, x*_7_ := *C*_*A*_*/N, x*_8_ := *C*_*U*_ */N* and *x*_9_ := *D/N* and *x*_10_ := *C*_*R*_*/N* hence state vector *<u>x</u>* := (*x*_1_, …, *x*_10_)^*tr*^, giving the dynamics for the probabilities *p*(*<u>x</u>, t*) as

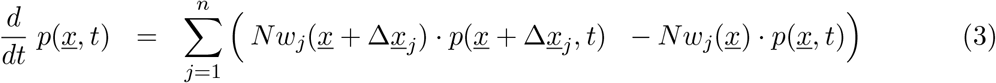

with *n* = 10 different transitions *w*_*j*_(*<u>x</u>*), as described by the mechanisms above, and small deviation from state *<u>x</u>* as 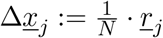 [44, 45, *40]*.*For the basic SHARUCD model we have explicitly the following transitions w*_*j*_ (*<u>x</u>*) and its shifting vectors *<u>r</u>*_*j*_ given by

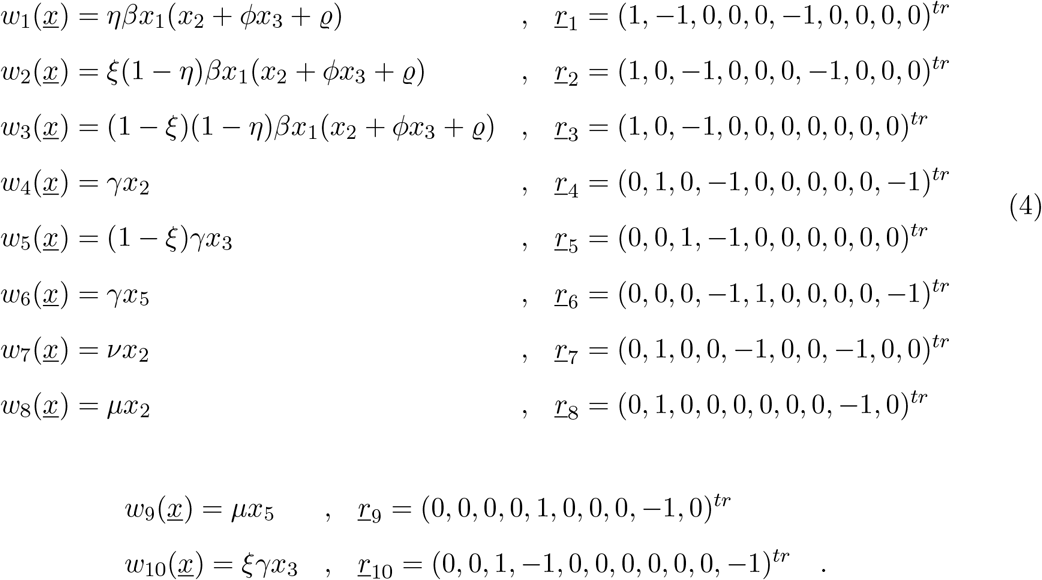

With these *w*_*j*_ (*<u>x</u>*) and *<u>r</u>*_*j*_ specified we also can express the mean field ODE system as shown in Appendix A.

### 2.3 Model simulations and data

Considering as starting points the biological aspects of the disease [27, 28, 30, 31, 32, 33], the model is calibrated and parametrized using the cumulative empirical COVID-19 incidence data for *C*_*H*_, *C*_*U*_ and *D* directly on the respective data sets, and for the positive tested infected *I*_*cum*_, which includes the new reported cases in all recording classes, except the recovered.

Parameters are estimated and fixed as the model is able to describe the disease incidence during the exponential phase of the outbreak, see Table 1 in Appendix A. The stochastic realizations of the model are calculated via the Gillespie algorithm [37, 38, 29], which is considered an exact algorithm once the governing equation, the master equation, is specified. Figure 2 shows the ensemble of stochastic realizations of the SHARUCD-type model, starting from March 4, 2020 until April 4, 2020. The period in time where the empirical data can no longer be described by the model simulations without control refers to the end of the exponential phase of the epidemic where the exponential growth decelerates into a growth close to zero towards a linear phase.

**Table 1:** Model parameters and initial condition values.

| Parameters,<br>variables and<br>initial conditions | Description | Values |
| --- | --- | --- |
| $N$ | population size | $2.2 \times 10^6$ |
| $H(t_0)$ | severe disease and hospitalized | 54.0 |
| $A(t_0)$ | mild disease and asymptomatic | 80.0 |
| $U(t_0)$ | ICU patients | 10.0 |
| $R(t_0)$ | recovered | 1.0 |
| $C_H(t_0)$ | recorded $H(t_0)$ | 54.0 |
| $C_A(t_0)$ | recorded $A(t_0)$ | 40.0 |
| $C_U(t_0)$ | recorded $U(t_0)$ | 10.0 |
| $C_R(t_0)$ | recorded $R(t_0)$ | 1.0 |
| $D(t_0)$ | death | 1.0 |
| $\beta$ | infection rate | $3.25 \cdot \gamma$ |
| $\phi$ | ratio of mild/asymptomatic infections<br>contributing to force of infection | $1.6[1.0 - 2.0]$ |
| $\gamma$ | recovery rate | $0.05d^{-1}$ |
| $\mu$ | disease induced death rate | $0.025d^{-1}[0.02 - 0.03d^{-1}]$ |
| $\nu$ | hospitalized to ICU rate | $0.025d^{-1}[0.025 - 0.1d^{-1}]$ |
| $\eta$ | proportion of hospitalization | $0.45[0.0 - 0.5]$ |
| $\xi$ | detection ratio of mild/asymptomatic | $0.45[0.0 - 0.8]$ |
| $\rho$ | import parameter | — |

**Figure 2:**
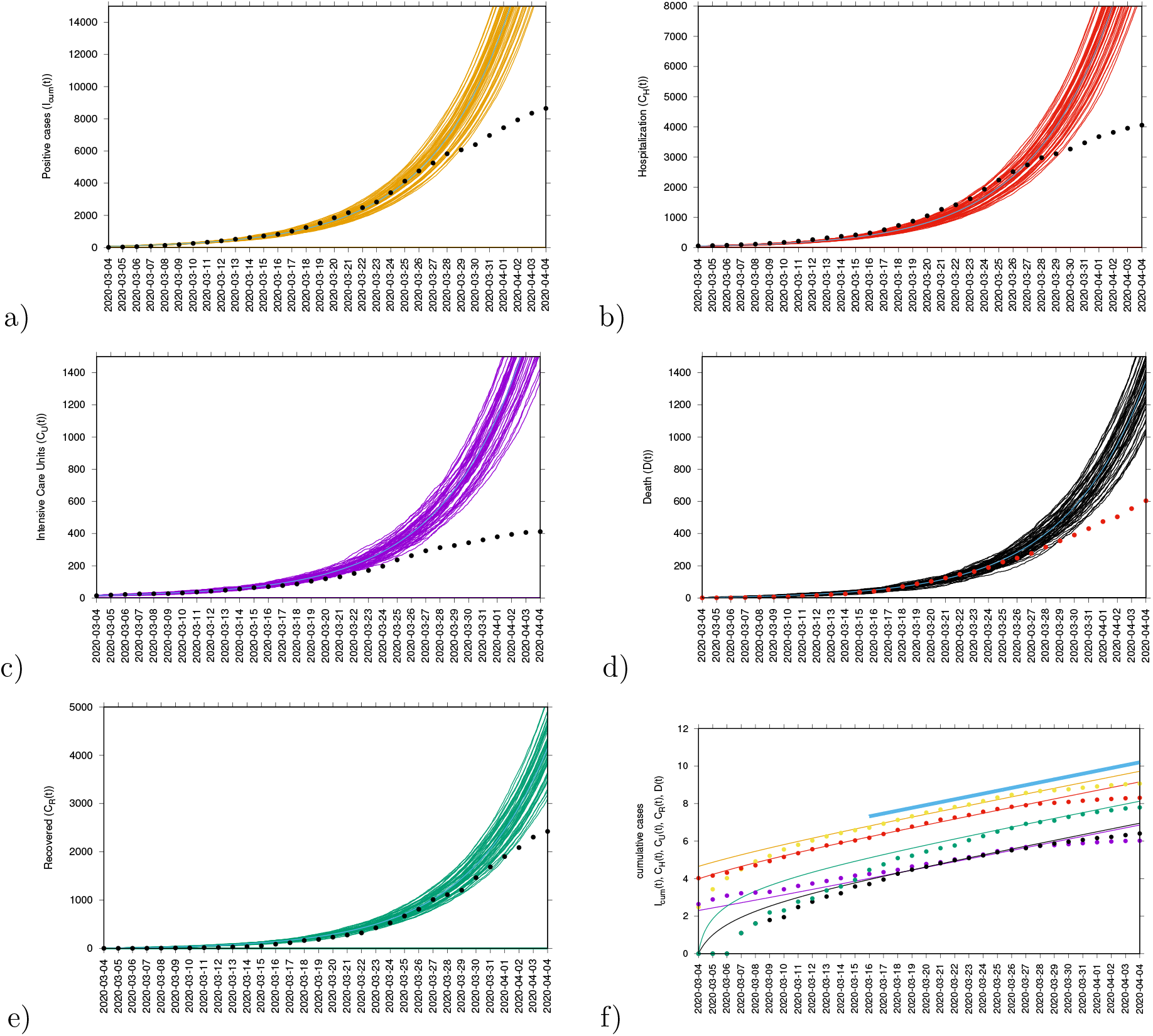
Ensemble of stochastic realizations of the SHARUCD-type model.The mean field solution is shown in light blue. a) Cumulative tested positive cases *I*_*cum*_(*t*), b) cumulative hospitalized cases *C*_*H*_(*t*), c) cumulative ICU admission *C*_*U*_ (*t*), d) cumulative deceased cases *D*(*t*), e) cumulative recorded recovered *C*_*R*_(*t*). In f) semi-logarithmic plot of the data and the mean field curves of all variables. For quite some time all mean field curves and the data are in parallel, and we could calculate the slope from the model parameters as growth rate *λ*, see light blue line.

Our model is able to describe the dynamics observed for each dynamical class, including the observed recovered (*C*_*R*_), for which data were only later available. This class was not used to determine the parameters, however, the data on the recorded recovered *C*_*R*_, including alive hospital and ICU discharges only, was immediately described, following the behavior observed from the other classes and calculating from the previously already estimated recovery rate *γ*. Figure 2 shows the ensemble of stochastic realizations of the SHARUCD-model and data.

### 2.4 Parameter uncertainties and model limitations

To investigate the parameter insecurities, we calculate numerically likelihood functions [41] for each parameter conditioned on the others and the data, with distances between simulations and data from all 5 variables, *D*(*t*), *I*_*cum*_(*t*), *C*_*H*_(*t*), *C*_*U*_(*t*) and *C*_*R*_(*t*), evaluated for the exponential phase of the epidemic. The likelihood plots for each individual parameter, recovery rate (*γ*), infection rate (*β*), disease induced mortality rate (*µ*), rate of ICU admissions (*ν*) and ratio of hospital admission due disease severity (*η*), the difference in infectivity of asymptomatic and hospitalized (*ϕ*) and the detection ratio of mild/asymptomatic infections (*ξ*) are shown in Fig. 3. Good agreement of the local maxima of the likelihood functions are obtained. The basic epidemiological parameters are shown in Fig. 3 a) to d) and the more internal ones, concerning the differences between mild and severe disease, are shown in Fig. 3 e) to g).

**Figure 3:**
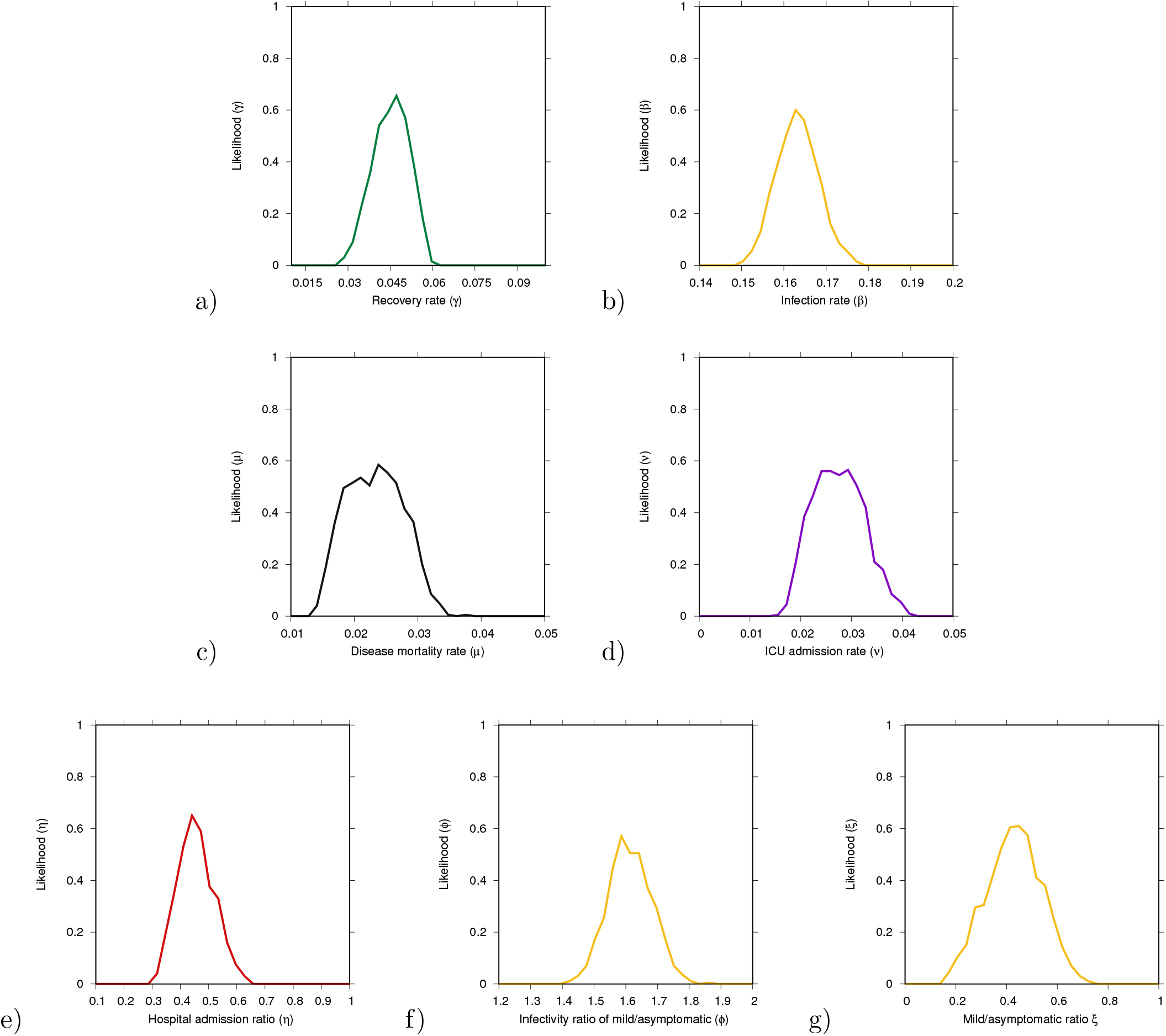
Numerical likelihood functions for the parameters a) recovery rate *γ*, b) infection rate *β*, c) diseased induced mortality rate *µ* and d) rate of ICU facilities admission *ν*, and e) hospital admission ratio *η*, f) infectivity of mild/asymptomatic relative to the hospitalized *ϕ* and g) recording rate of mild/asymptomatic cases *ξ*.

Although the parameter set used to describe the exponential phase of COVID-19 outbreak in the Basque Country are coinciding well with the calculated maximum likelihood values, parameters are prone to correlations, often only determined as combinations but not individually on scarce data. Possible correlations are investigated in Appendix B. Models output are based on the data available, which are often incomplete, and for non-existing data such as data referring to the proportion of undetected asymptomatic infected as well as how infective those individuals are (i.e. their contribution to the force of infection as compared to the symptomatic detected individuals) during the exponential phase of the epidemic are estimated but not yet validated with empirical data. However, these data would eventually change the dynamical behavior obtained for the positive cases and recovered and for the other variables it would remain the same. We keep calibrating the model framework with updated data and so far, the selected parameter set is still used also during the control phase without need of adjustments.

### 2.5 Modeling the effects of the control measures

With the initial parameters estimated and fixed on the exponential phase of the epidemic, we model the effect of the disease control measures introduced using a standard sigmoid function *σ*(*x*) = 1*/*(1 + *e*^*x*^), shown along the main results in Fig. 6 b) (black line), which is able to describe well the gradual slowing down of the epidemics, as it turned out later in the response of the disease curves to the control measures, see Fig. 6 and its detailed description below. Specifically, the infection rate *β* becomes time dependent with *β*(*t*) = *β*_0_*σ*_−_(*x*(*t*))+*β*_1_*σ*_+_(*x*(*t*)), where *σ*_−_ := 1*/*1 + *e*^*x*^ is a downward sigmoidal and *σ*_+_ := 1*/*1 + *e*^−*x*^ is an upward sigmoidal. The time depend argument is *x*(*t*) := *a*(*t* − *t*_*c*_). Modifications of the lower value of infectivity *β*_1_ might be needed when the relaxation of the control measures can change the contact probabilities again or in the eventual case of seasonality as observed in other respiratory diseases.

#### 2.5.1 Short-term prediction exercise with control measure

Short-term predictions considering the effective control measures described above are shown in Fig. 4. For this exercise, empirical data were available up to April 13, 2020, and model simulations are obtained for seven days longer run than the available data (see Fig. 4 a, c, e, g, i). A week later, new data were included to check the quality of the short prediction exercise (see Fig. 4 b, d, f, h, j). The mean field solution without the control function is plotted in light blue, indicating the differences of model prediction with and without control measures. In good agreement, hospitalization and deceased cases are well matched within 50 stochastic simulations obtained with the Gillespie algorithm, with data lying in the median range of stochastic realizations. Note that for the likelihood functions we use several hundreds of stochastic realizations. The ICU admission data look atypical and can not be described with the current control scenario, after the exponential phase of the epidemic. This aspect is investigated in more detail in the following sections, using further measures and model refinements.

**Figure 4:**
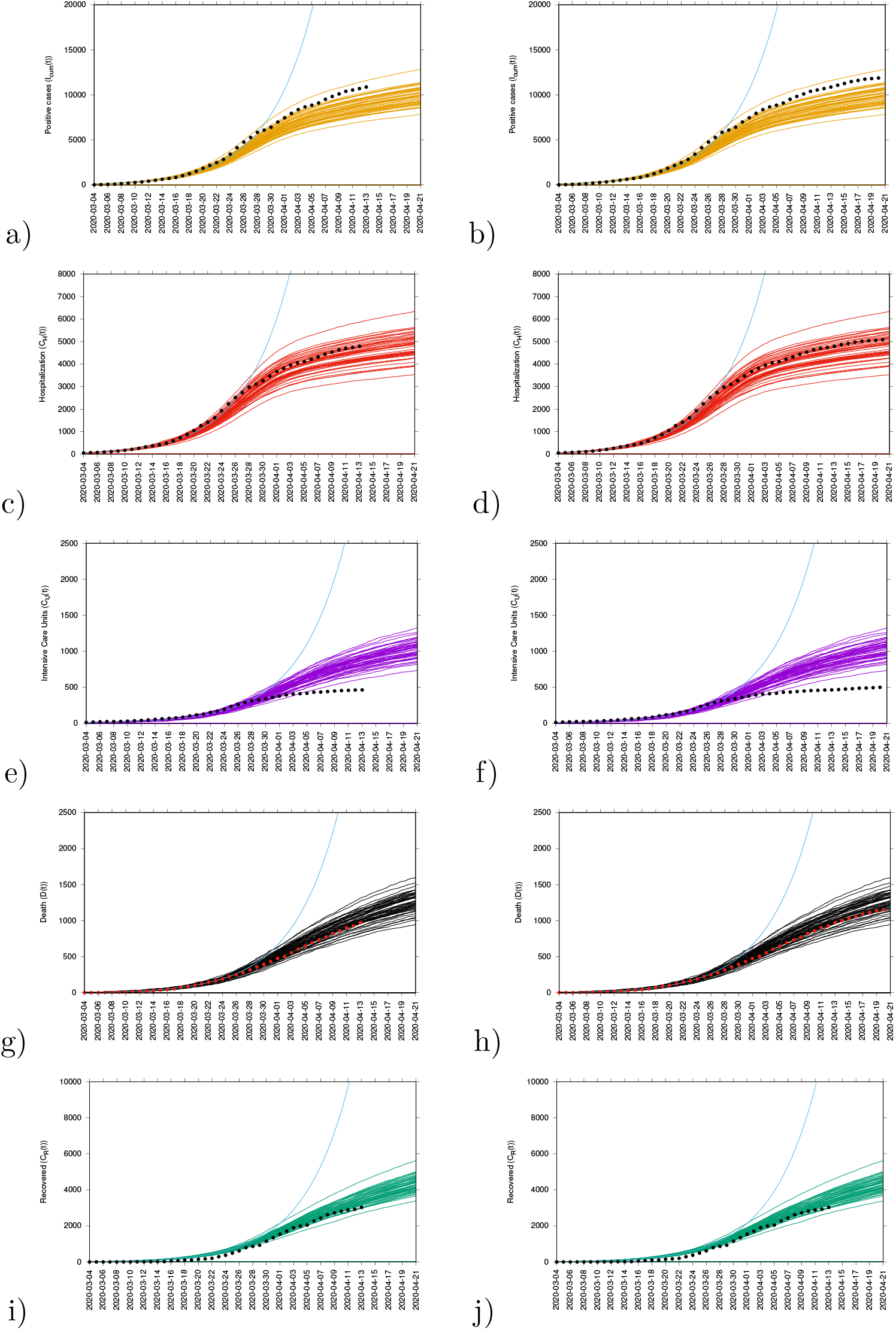
Ensemble of stochastic realizations of the SHARUCD-model with control and data matching, starting from March 4, 2020. a-b) Cumulative tested positive cases *I*_*cum*_(*t*), c-d) cumulative hospitalized cases *C*_*U*_ (*t*), e-f) cumulative ICU admission *C*_*U*_ (*t*), g-h) cumulative deceases cases *D*(*t*), i-j) cumulative recovered *C*_*R*_(*t*).

The cumulative incidences for tested positive cases (*I*_*cum*_) follow the higher realizations range whereas the cumulative incidences for alive hospital discharges (*C*_*R*_), a proxy for notified recovered individuals which were hospitalized because of COVID-19, but not including the recovered individuals which were eventually tested positive but did not need hospitalization, follow the lower realizations range. While the deviation observed for the total tested positive (*I*_*cum*_) can be explained by the increased testing capacities over time since March 22, 2020, where more cases are expected to be detected, including sub-clinical infections and eventually asymptomatic individuals, the deviation observed for the “recovered” individuals are as expected, since the dynamical variable *C*_*R*_ counts, besides the notified *H* + *U* alive discharges, a proportion of tested positive mild/asymptomatic individuals that did not need hospitalization (*ξA*).

#### 2.5.2 Long-term prediction with control measures

Longer-term predictions for hospital admission and deceased cases are obtained, with very small numbers of new hospital admission cases (close to zero increment) around 130 days after March 4th, 2020. Deceased cases are predicted to reach zero increment 2-3 weeks later, due to the delay between the onset of symptoms, hospitalization and death. The mean numbers are estimations where wide confidence intervals should be taken into as visible from the the higher ranges of model realizations. Data used in this exercise were not discriminated by diagnostic method and therefore the final numbers might contain some overlap and possible double notification occurring when the rapid tests were introduced as screening tool in the Basque Country.

### 2.6 Growth rate and reproduction ratio

After an introductory phase, the epidemic entered into an exponential growth phase, which started in the Basque Country around March 10, 2020, and due to the effects of the imposed control measures has left the exponential growth phase to a slower growth around March 27, 2020, see Fig. 2.

The exponential growth phase is typical for any outbreak with disease spreading in a completely susceptible population, as observed already in the SIR-system, Eq. (1), with 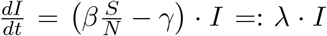 for *S* ≈ *N* and hence 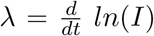 For the present SHARUCD model we obtain similarly an exponential growth factor analytically. From the active disease classes *H* and *A* with the dynamics given by

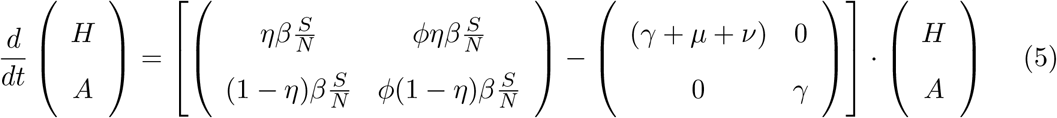

we obtain 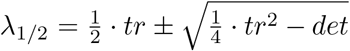 with the parameter dependent trace *tr* = (*η* + *ϕ*(1 − *η*)) *· β* − (2*γ* + *µ* + *ν*) and determinant *det* = *γ*(*γ* + *µ* + *ν*) − ((*γ* + *µ* + *ν*)*ϕ*(1 − *η*) + *γη*) *· β*. The dominating growth factor is then given by the largest eigenvalue *λ*_1_. The concept of the growth rate can be extended into the phase when effects of the control measures become visible and parameters slowly change, such that for short times the above analysis holds as for constant parameters.

Another measure of the spreading of the disease in its initial phase is the basic reproduction number (*R*_0_), the number of secondary cases *I*_*s*_ from a primary case *I*_*p*_ during its infectivness before recovering in a completely susceptible population, giving in the SIR dynamics reproduction ratio *r* = *β/γ*. From the next generation matrix in the case of the SHARUCD model we obtain the dominant eigenvalue 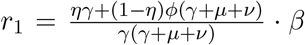 as the reproduction ratio in a completely susceptible population. This concept can be also extended to larger compartmental models and into the phase when effects of the control measures become visible and parameters slowly change. The momentary reproduction ratios *r* can be analyzed, as frequently done for the COVID-19 epidemics, but often called “basic reproduction number”. While the momentary growth rate follows directly from the time continuous data at hand, the momentary reproduction ratio depends on the notion of a generation time *γ*^−1^. The momentary growth rates and momentary reproduction ratio are analyzed below.

#### 2.6.1 Momentary growth rates and momentary reproduction ratio

We calculate the growth rates and reproduction numbers for the Basque Country from the COVID-19 data at hand available, from March 4 to May 4, 2020. While we do not take responsibility for the absolute value of *r*(*t*) as it is bound to many internal assumptions, recovery period, smoothing and approximations, which are not valid for time dependent parameters, the threshold behavior is independent of those uncertainties and clearly indicates that the outbreak is under control at this very moment, allowing a gradual lockdown lifting restrictions under constant monitoring.

To obtain the momentary growth rates from data directly we use 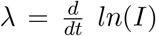 at first applied to the cumulative tested positive cases *I*_*cum*_(*t*) obtaining, via a smoothing window, the new cases after time *τ* As

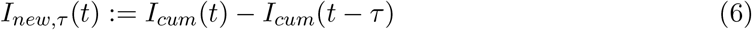

and hence, the growth rate

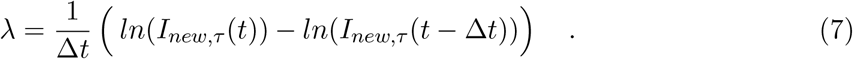

From the growth rate, the reproduction ratio is calculated with the recovery period *γ*^−1^ obtained from underlying models and recent literature about SARS-CoV-2 interaction with human hosts [27, 28, 30, 31, 32, 33]. The results are given as data dots in Fig. 6 a) for the growth rates and in b) for the reproduction ratios and the black curves from the control response *β*(*t*) with its sigmoidal shape using the SHARUCD-model expressions *λ*_1_ in a) and *r*_1_ in b). After an original introductory phase of the epidemic where insecurities in the data collection (due to small numbers) were still present while setting up the recording system, the curves agree well, from around March 14, 2020 on, with surprisingly good results, also in the lower value of the sigmoidal curve.

**Figure 5:**
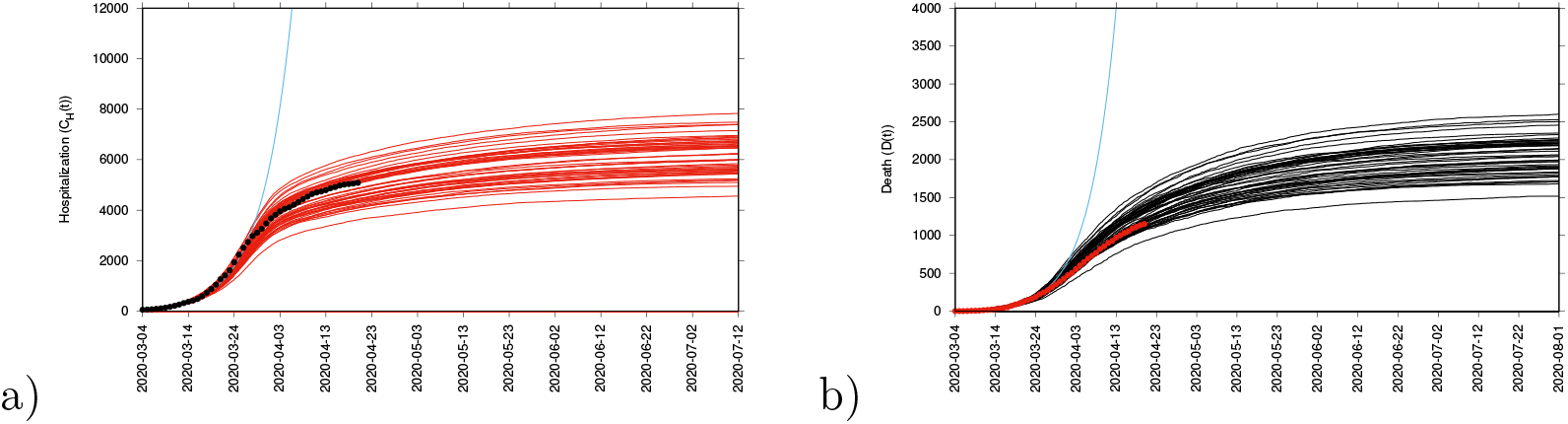
Ensemble of stochastic realizations of the SHARUCD-model, a) cumulative hospitalized cases *C*_*H*_(*t*) and b) cumulative deceases cases *D*(*t*) for long term predictions

**Figure 6:**
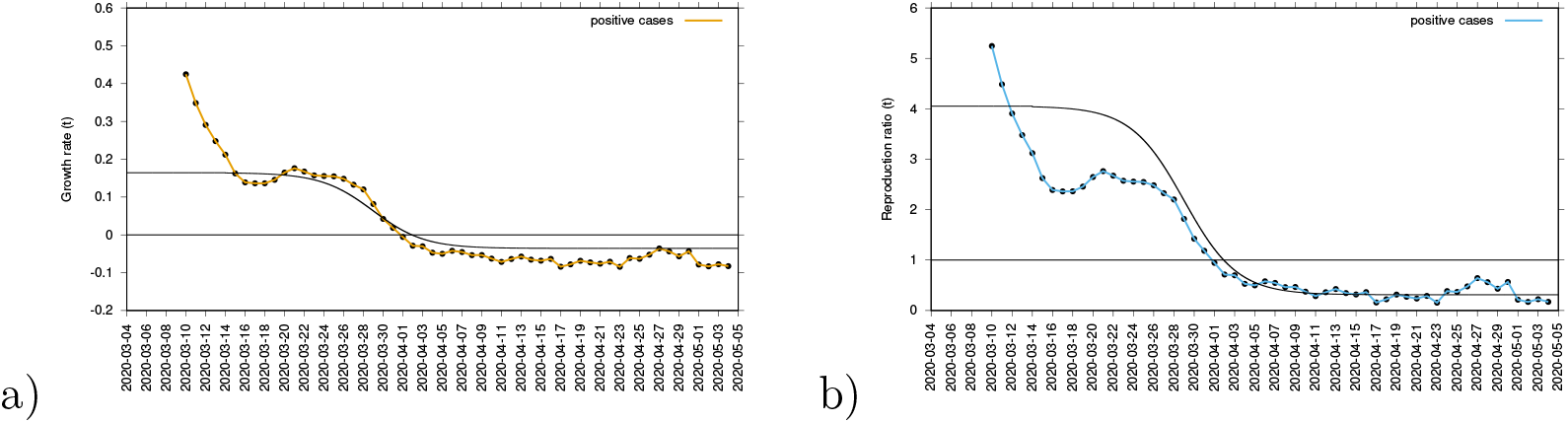
a) Growth rate estimation from the data on positive tested infected cases, and b) reproduction ratio from the same data.

The concept of growth rates can be extended to the other measured variables, hospitalizations, deceased cases, recovered cases and ICU admitted cases, see Fig. 7. The sigmoidal shape of decreasing growth rates is well visible in the hospitalized and the tested positive infected, whereas the deceased and the recovered are following only later with a delay of about 8 to 10 days, and a much slower sigmoidal curve or near to linear decline. Notably, the ICU admitted cases follow the sharp sigmoidal decline of growth rate of hospitalized and tested positive cases rather than the growth rate of deceased and the recovered. These results let to information about how to refine the model in order to capture the dynamics of the ICU admissions, better than in the present model as we will describe in the next subsection.

**Figure 7:**
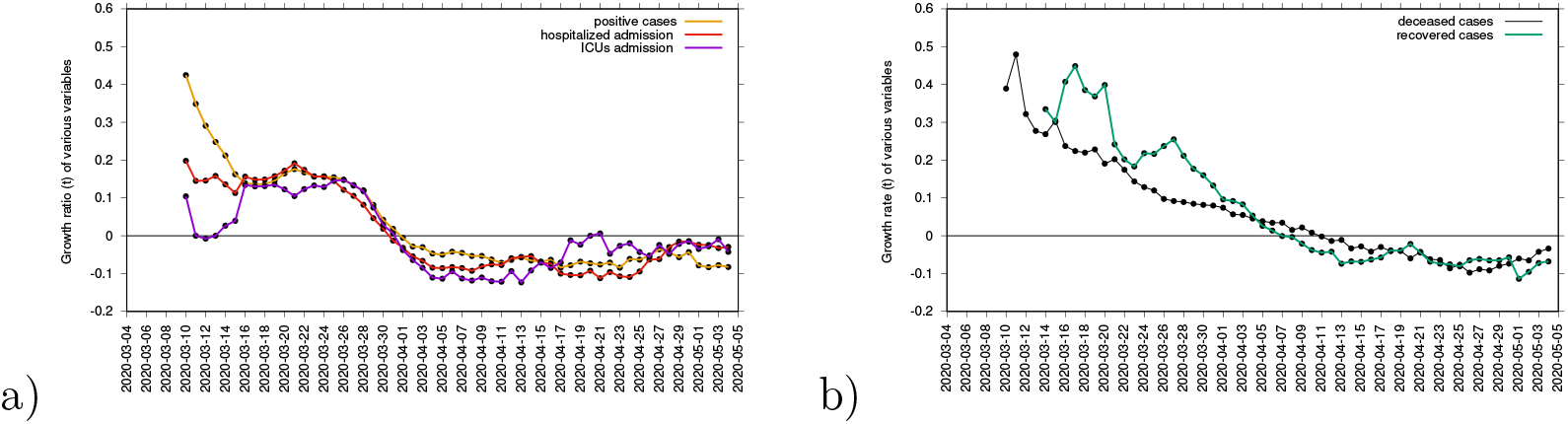
Growth rate estimation for various variables. In a) growth rate for tested positive cases (yellow), hospitalizations (red) and ICU (purple). In b) growth rate for recovered (green) and deceased (black). Clearly, the two groups can be distinguished.

### 2.7 Model refinement based on observations from growth rates of the different data sets

Given the observed synchronization of the ICU admission cases with the cumulative tested positive cases and hospitalizations, the SHARUCD model is now refined, using data available from March 4 to May 4, 2020. We change the transition into ICU admissions from the previous assumption of hospitalized patients recovering with recovery rate *γ*, being admitted to ICU facilities with rate *ν* or dying with disease induced death rate *µ* by the assumption that ICU admissions are consequence of disease severity prone to hospitalization analogously to rate *η*.

The updated transitions are changed from the previously used form

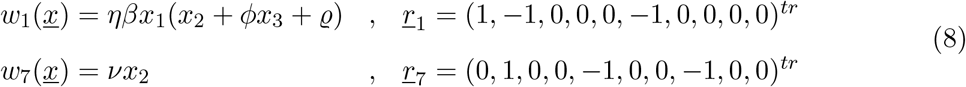

into

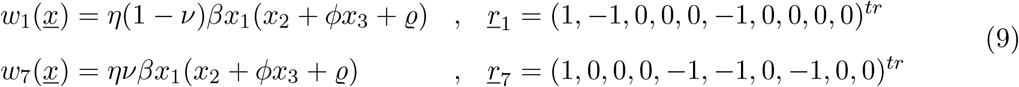

with the parameter *ν* adjusted from the ICU-admission rate in units of *d*^−1^ into an ICU- admission ratio *ν* ∈ [0, 1]. By changing and fixing *ν* = 0.1, we obtain immediately a very good agreement between the available cumulative ICU data and model simulations, with only small deviations in the other variables, see Fig. 8 c).

**Figure 8:**
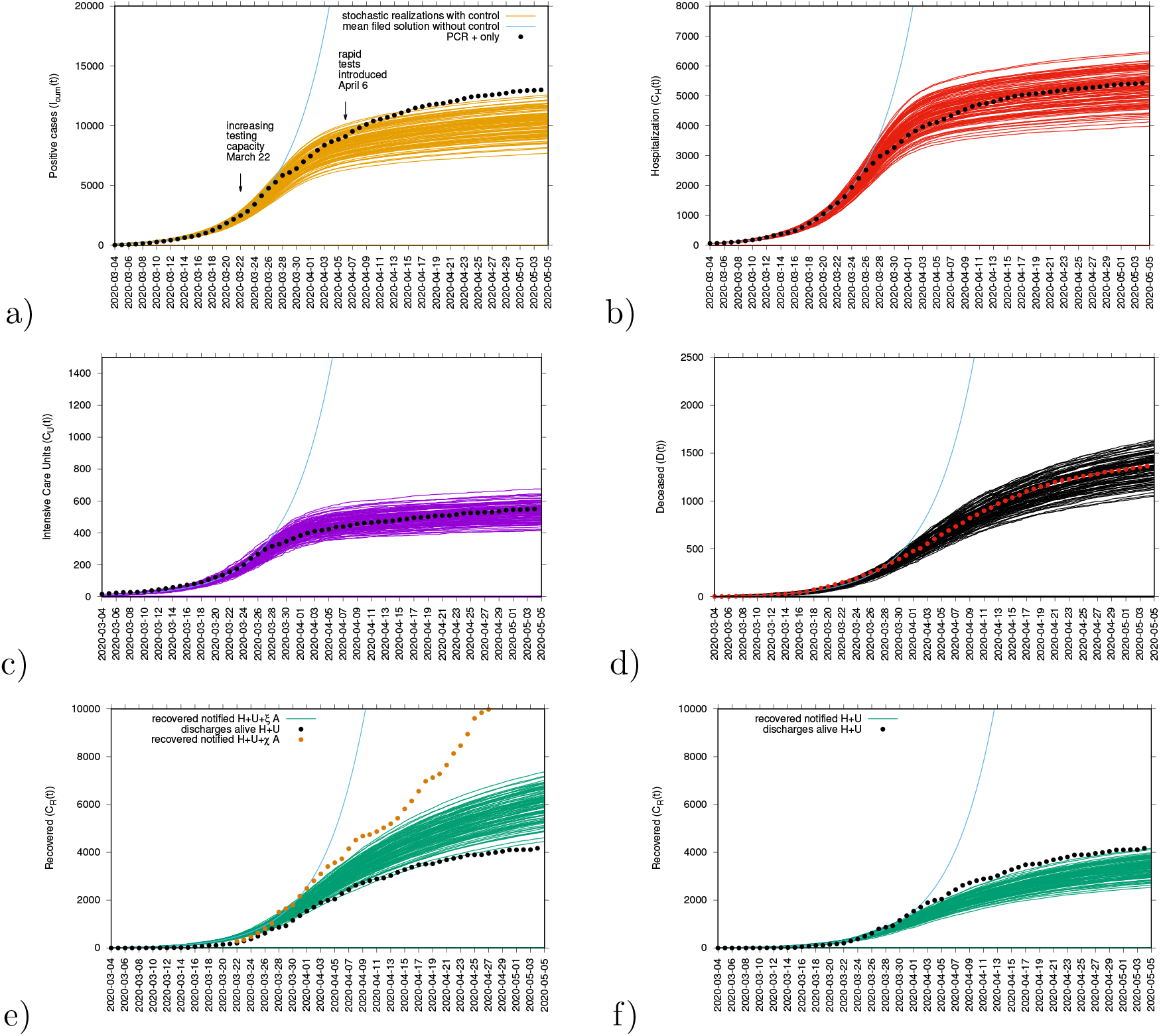
Ensemble of stochastic realizations of the refined SHARUCD-model and data matching. The mean field solution without control is shown in light blue. a) Cumulative tested positive cases *I*_*cum*_(*t*), b) cumulative hospitalized cases *C*_*U*_ (*t*), c) cumulative ICU admissions *C*_*U*_ (*t*), d) cumulative deceases cases *D*(*t*), e-f) cumulative recovered *C*_*R*_(*t*).

In good agreement, the refined model can describe well the hospitalizations (see Fig. 8 b)), the ICU admissions and the deceased cases (see Fig. 8 d)), well matched within the median of the 100 stochastic realizations from the model. The cumulative incidences for tested positive cases (RT-PCR+ only) still follow the higher realizations range (see Fig. 8 a)), due to the increasing testing capacities since March 22, 2020, followed by the introduction of rapid tests as screening tool that the current model (without testing feedback) can not describe quantitatively. The cumulative incidences for alive hospital discharges (black dots), used as a proxy for notified recovered individuals needing hospitalization because of COVID-19 only, keeps following the lower realizations range, when model simulations are obtained for the cumulative notified recovered (*H* + *U* + *ξA*), shown in Fig. 8 e). Orange dots refers to all notified recovered (*H* + *U* + *χA*), including also tested positive mild/asymptomatic infections detected when the testing capacity increased. To describe this new data set, a new transition is needed to record the proportion of recovered from detected mild/asymptomatic infections (*χ*). Figure 4 8 f) shows model simulations obtained for the cumulative notified recovered from hospital (*H* + *U*) only. Similarly, in order to describe quantitatively this specific available data set, a new transition is needed to record a proportion of recovered individuals tested positive without being admitted to the hospital (*ξA*).

The calculations of the growth factors and reproduction ratios were updated to the new model modifications, namely from the disease class dynamics now as

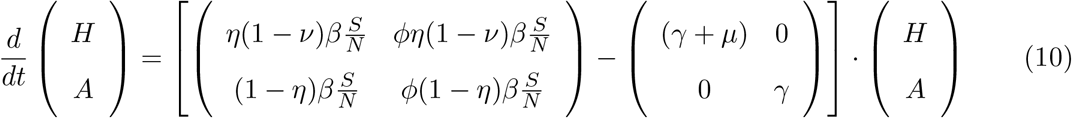

we obtain *λ*_1_ via 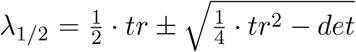 with *tr* = (*η*(1 − *ν*) + *ϕ*(1 − *η*)) *· β* − (2*γ* + *µ*) and *det* = *γ*(*γ* + *µ*) − ((*γ* + *µ*)*ϕ*(1 − *η*) + *γη*(1 − *ν*)) *· β* and the reproduction ratio as 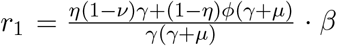 From Fig. 9 we observe the data to follow the exponential growth phase of COVID-19 epidemic in the Basque Country from about March 14 to March 27, 2020, sensing the slow down due to the control measures from March 28, 2020 onwards. Fig. 9 b) shows the semi-logarithmic plot for the refined model and its adjusted parameter set matching with the data for all variables. The analytically calculated growth rate *λ*_1_ (light blue line) is also shown, close to the mean field solutions (parallel straight lines) for the exponential phase of the outbreak.

**Figure 9:**
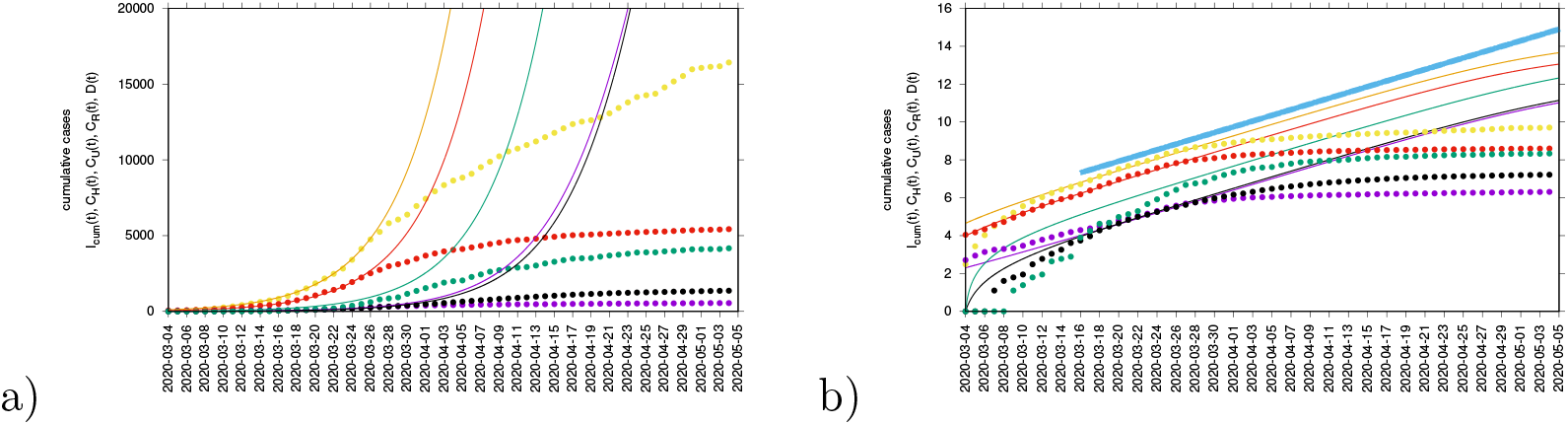
Adjusted SHARUCD model to synchronize ICU admissions with tested positive cases and hospitalizations. In a) data and mean field solutions in natural scale. In b) data and mean field solutions in semi-log scale with adjusted mean field curves and growth rate *λ* as light blue line.

**Figure 10:**
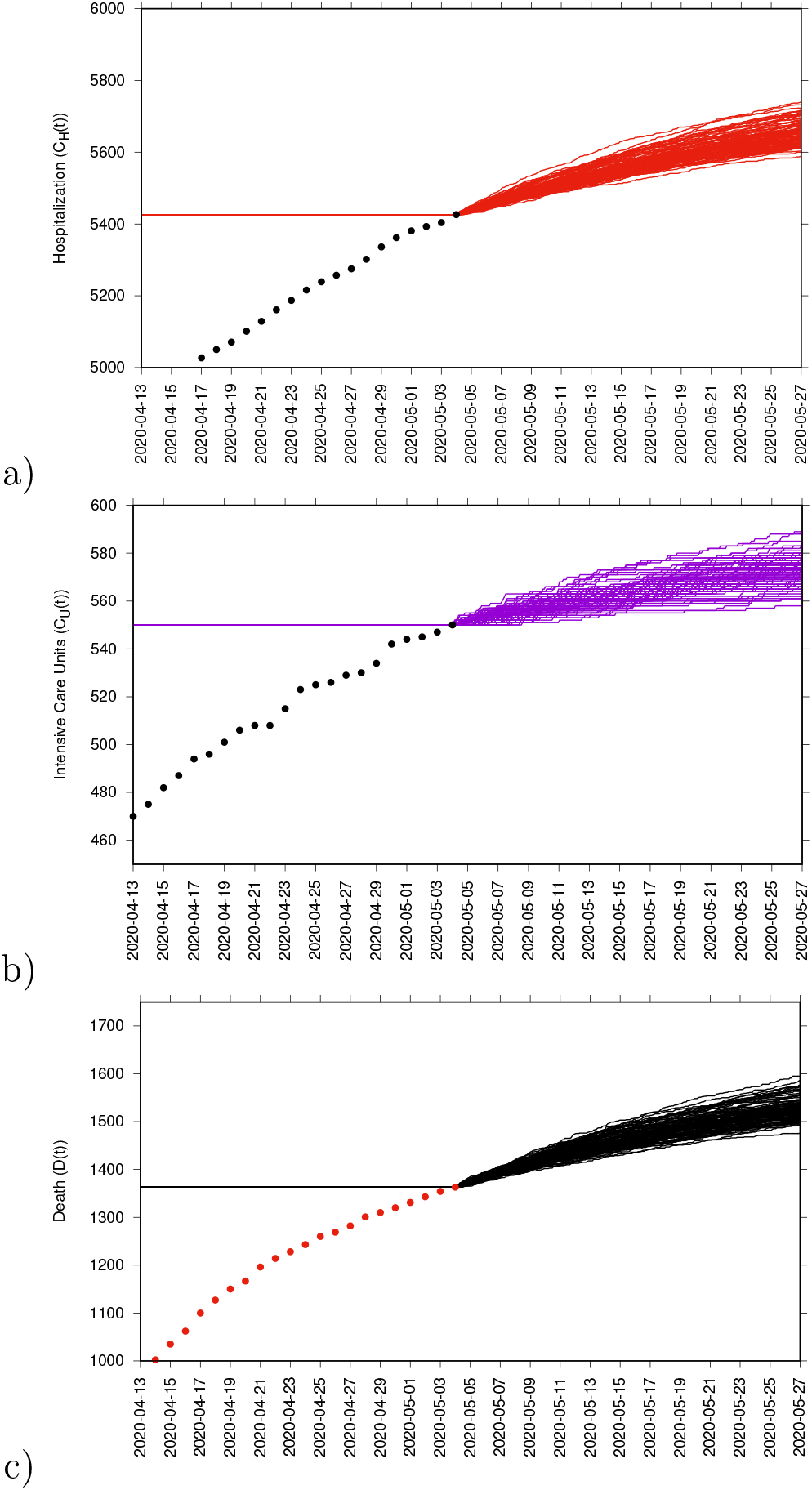
a) Cumulative hospitalized cases *C*_*H*_(*t*), b) cumulative ICU admissions *C*_*U*_ (*t*) c) cumulative deceased cases *D*(*t*).

#### 2.7.1 Prediction up to May 25th

Medium-term prediction exercise under constant external conditions was performed using the last data point available on May 4, 2020. Taking the minimum and the maximum ranges of the stochastic realizations as reference, the number of new hospitalization is predicted to be between *≈* 5600 to *≈* 5750 cumulative cases up to May 25, 2020 and ICU admissions between *≈* 560 to *≈* 590 cumulative cases up to May 25, 2020. The number of deceased is predicted to be between *≈* 1500 to *≈* 1600 by May 25th, referring to the number of predicted confirmed severe cases from 2-3 weeks before (May 5th), due to the delay between the onset of symptoms, hospitalization and death.

## 3 Discussion and future work

In March 2020, a multidisciplinary task force (so-called Basque Modelling Task Force, BMTF) was created to assist the Basque Health managers and the Basque Government during the COVID-19 responses. In this paper we analyze the results obtained with a stochastic SHARUCD-type model approach, taking into consideration all information provided by the public health frontline and keeping the biological parameters for COVID-19 in the range of the recent research findings, but adjusting to the phenomenological data description. Using the data provided by the Basque Health Service, models were able to describe the disease incidence data with a single parameter set.

A careful data inspection has shown the end of the exponential phase of the epidemic to be around March 26, 2020, allowing us to infer that the partial lockdown was effective and enough to decrease disease transmission in the Basque Country. Disease control was modeled, given the current epidemiological scenario, able to describe well the gradual slowing down of the COVID-19 outbreak. Short-term prediction exercises were performed, with seven days longer simulation runs than the available data, where hospitalizations, recovered and deceased cases matched well within the median of the stochastic model simulations. The ICU admission data looked atypical under the baseline proposed model and could only be described qualitatively during the exponential phase of the outbreak, however not well quantified afterwards. The cumulative incidence for all positive cases followed the higher stochastic realizations range instead and the deviation observed between model simulations and data was interpreted by the increased testing capacities with not only the expected increment in the number of new positive cases, including sub-clinical infections and eventually asymptomatic individuals, but also with some overlap and possible double notification occurring when the rapid tests were introduced as screening tool in nursing homes and for the frontline public health workers.

Growth rates (*λ*(*t*)) and the reproduction ratio (*r*) were calculated from the model and from the data and the momentary ratio *r* is estimated to be below the threshold behavior of *r* = 1, but still close to 1, meaning that although the number of new cases reported in the Basque Country are decelerating, the outbreak is still in its linear phase and careful monitoring of the development of the dynamics of the new cases from all variables and respectively all data sets is required. The growth rates for various variables are negative, confirming the momentary decrease in disease transmission. Moreover, we observe that the dynamics of the hospitalizations and ICU admissions were synchronized with the total tested positive cases, becoming negative on April 1, 2020, whereas recovered and deceased cases only follow later, reaching negative growth rate on April 7 and April 11, 2020, respectively. These results lead to a model refinement to synchronize ICU admissions to hospitalizations and to positive tested infected, rather than to deceased and to recovered. The refined model is now able to describe well the 5 variables, with the empirical data lying in the median range of stochastic realizations using a single parameter set.

As the concept of *R*_0_ used alone can be easily misinterpreted, specially now when testing capacity is increasing, with momentary reproduction ratios expected to be influenced though to a gradual augmentation, leading to a rather misleading indication that the epidemic is no longer “under control”, as it is bound to many internal model assumptions, smoothing and approximations, significantly affected by the assumed recovery period. The BMTF is monitoring the development of the COVID-19 epidemic in the Basque Country by also evaluating the momentary growth rates *λ*(*t*) for the positive cases *I*_*cum*_(*t*), but also the *λ*(*t*) for hospitalizations (*C*_*H*_), ICU admissions (*C*_*U*_), deceased (*D*) and recovered cases (*C*_*R*_). Without interfering in any political decision, we assist the Basque Health Managers and the Basque Government with results that are obtained by this model framework, based on available data and evidence as scientific advice.

A mid-term prediction exercise is shown, under constant external conditions, with a note of wide confidence intervals considering the higher ranges of model realizations. Models limitations and the implication of using different available data sets are discussed. As future work, a slight adjustment of the model could further improve the description of the tested positive cases dynamics and the recovered via testing feedback. This will become more important in the future course of the epidemic and will give us better information on the level of asymptomatic and mild infections, allowing to infer on population immunity development in the course of the year and eventually the following years.

## Data Availability

Epidemiological data used in this study are provided by the Basque Health Department and the Basque Health Service (Osakidetza), continually collected with specific inclusion and exclusion criteria.

https://www.euskadi.eus/boletin-de-datos-sobre-la-evolucion-del-coronavirus/web01-a2korona/es/

## Acknowledgements

Maíra Aguiar has received funding from the European Union’s Horizon 2020 research and innovation programme under the Marie Skodowska-Curie grant agreement No 792494. Data were provided by the Basque Health Department and the Basque Health Service (Osakidetza). We thank the huge efforts of the whole COVID-19 BMTF, specially to Adolfo Morais Ezquerro, Vice Minister of Universities and Research of the Basque Goverment, and Jose Antonio Lozano, Scientific Director of BCAM for the fruitful discussions.

## Author contributions statement

M.A and N.S. have conceived of the study, developed and analyzed the models. E. M. has collected and prepared the data sets used in this study. M.A., E. M., N.S, J.B.V and J.M have contributed with manuscript writing and critical discussions. All authors reviewed the manuscript.

## Additional information

## Appendices

### A Mean field approximation and stochastic differential equation approximation of the basic SHARUCD model

From the master equation for state discrete stochastic processes, given in densities, Eq. (3), we derive now a stochastic differential equation system as diffusion approximation, which as by-product also gives the mean field approximation as deterministic drift term in the Fokker-Planck equation. We use Taylor’s expansion for small changes of densities Δ*<u>x</u>*_*j*_, hence

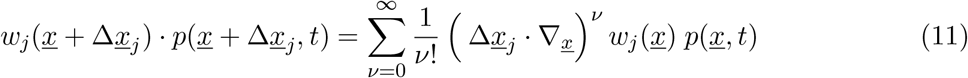

giving to second order in 1*/N* a Fokker-Planck equation

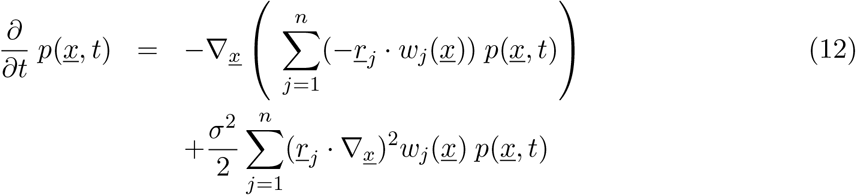

with 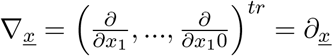 or in different notation

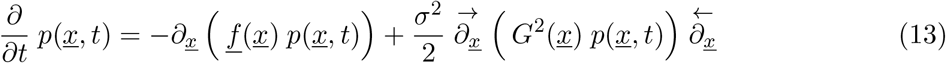

using simply a quadratic form 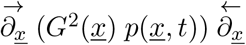 here with

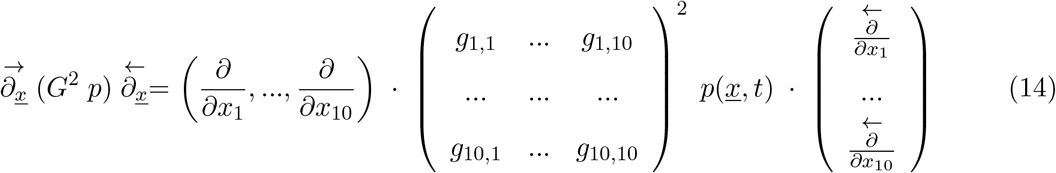

and

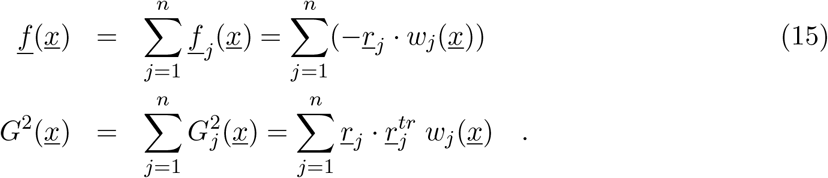

The Fokker-Planck equation gives a stochastic differential equation system with 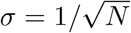 and Gaussian normal noise vector 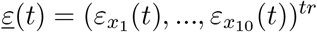 as

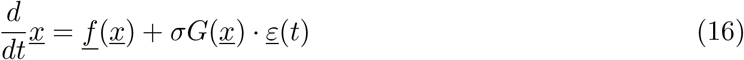

and using matrix square root from eigenvalue-eigenvector decomposition 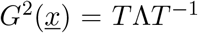 as 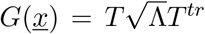 to be numerically implemented easily, and much faster than the Gillespie algorithm for the master equation, when it comes to large population sizes *N* and longer runs of e.g. one year, as for a complete disease outbreak curve necessary. For possibly non-quadratic matrices *B*, expressing the covariance matrix as *B*^*tr*^*B* = *G*^2^, see [39], speading up further the stochastic process simulations.

In mean field approximation we obtain explicitly from

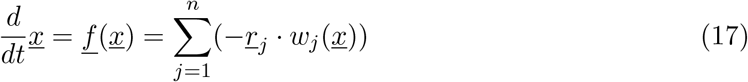

with the transitions *w*_*j*_(*<u>x</u>*)) specified in Eq. (5). The deterministic version of the model is given by a differential equation system for all classes, including the recording classes of cumulative cases *C*_*H*_, *C*_*A*_, *C*_*R*_ and *C*_*U*_ by

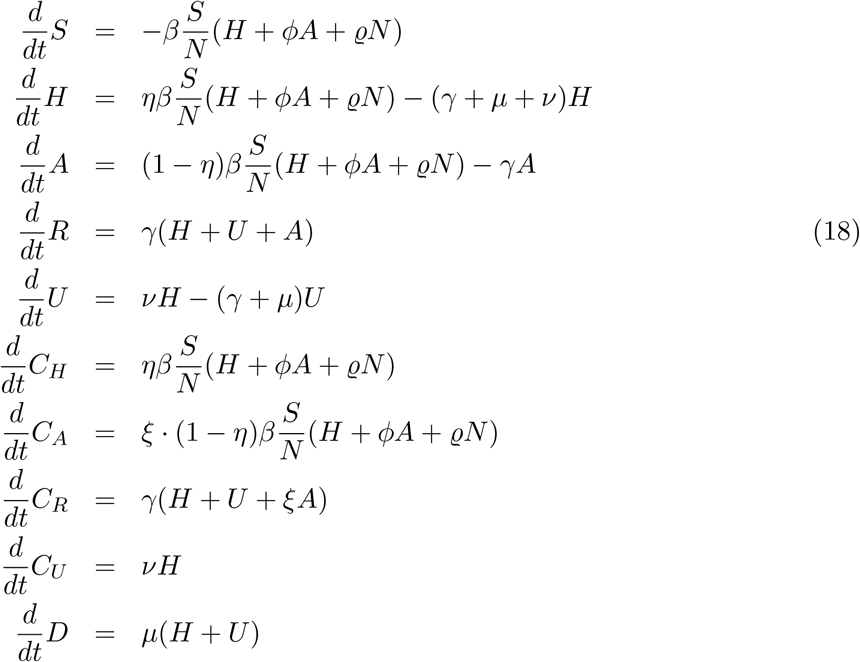

in a complete form.

For a constant population size *N*, susceptible individuals (*S*) become infected with SARS-CoV-2. With proportion *η*, individuals become severe cases requiring hospitalization (*H*), always detected via positive RT-PCR test or double testing with a first rapid test for antibodies, though up to now being quite unspecific, and a subsequent more specific RT-PCR test, whereas with proportion 1 − *η* individuals develop mild/asymptomatic infection, which are detected with a ratio *ξ* (∈ [0,1]). Severe hospitalized individuals transmit the disease with infection rate *β* whereas mild/asymptomatic cases transmit the disease with infection rate *ϕβ*. This difference in transmission is reasonable when assuming that, once hospitalized, individuals are isolated and would no longer transmit as much as mild/asymptomatic cases, often undetected and more mobile. Infected individuals may recover with a recovery rate *γ*, however, severe hospitalized individuals could be also admitted to the ICU facilities, with a rate *ν*, or die before being admitted to the ICU facilities, with disease induced death rate *µ*. Individuals admitted to the ICU facility could eventually recover with a recovery rate *γ* or die with disease induced death rate *µ*.

The model is calibrated using the empirical data for the Basque Country and the biological parameters are estimated and fixed as the model is able to describe the disease incidence during the exponential phase of the epidemic for each dynamical class. Parameter insecurities are quantified with likelihood functions. Model parameters and initial conditions are shown in Table 1, where *β* is the infection rate and *ϕ* is the ratio describing the asymptomatic/mild infections contribution to the force of infection. *γ* is the recovery rate, *µ* is the disease induced death rate and *ν* is the rate of hospitalized going to the ICU. *η* is the proportion of susceptible being infected, develop sever symptoms and being hospitalized whereas 1 − *η* is the proportion of susceptible becoming infected and developing mild disease or asymptomatic. *ξ* is the ratio of detected, via testing, mild/asymptomatic infect individuals. *ϱ* is the import rate needed to describe the introductory phase of the epidemics and for the present study, we assume *ϱ* to be much smaller than the other additive terms of the force of infection, given the strong observational insecurities on the data collected at the beginning of the outbreak.

#### A.1 The refined model

The original SHARUCD model was refined to synchronize ICU admissions to hospitalizations and positive tested infected, rather than to deceased and recovered. We consider primarily SHARUCD model versions as stochastic processes in order to compare with the available data which are often noisy and to include population fluctuations, since at times we have relatively low numbers of infected in the various classes.

We change the transition into ICU admissions from the form like used in recovery *γ* and death *µ*, more to the one for distinction of hospitalized and asymptomatics *η*, to describe the results from the growth rate analysis of all data sets, see previous subsection. Hence we update the transitions just a bit from

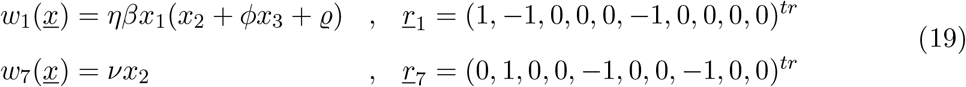

into

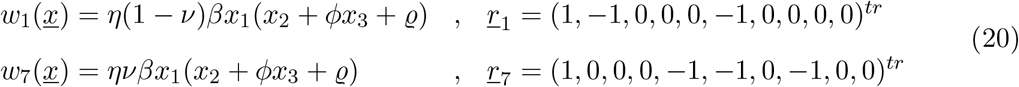

and have to adjust the parameter *ν*, which was an ICU-admission rate in units of *d*^−1^ into an ICU-admission ratio *ν* ∈ [0,1]. Parameters were slighly adjusted and for the refined model we use *ν* = 0.1, *µ* = 0.02, *ϕ* = 1.65, *ξ* = 0.4 and *η* = 0.4 and obtain immediately a very good agreement now of ICU data and simulations, with the empirical data for each variable lying in the median range of stochastic realizations, and only small deviations in the other variables. The other parameters values were kept the same as shown in Table 1.

### B Two parameter likelihood plots

We analyzed possible correlations between parameters by inspecting numerical two-parameter likelihood functions. Since the growth factor *λ* is mainly determined as *λ ≈ β* − *γ* we observe in many basic epidemiological models large correlations between especially these two parameters. Large infectivity *β* and small recovery period *γ*^−1^ describe often data as well as small infectivity and long recovery period. However we observe in a first inspection of the numerical likelihood of these two parameters, obtained from considering all 5 paremter sets in comparison with the SHARUCD basic model, that the range of possible values for *β* and *γ* together, see in Fig. 11 the area lifted away from zero in *L*(*β, γ*), is quite restricted around the best values for each of the parameters individually, as they are shown in Fig. 3 a) and b) in the main text.

**Figure 11:**
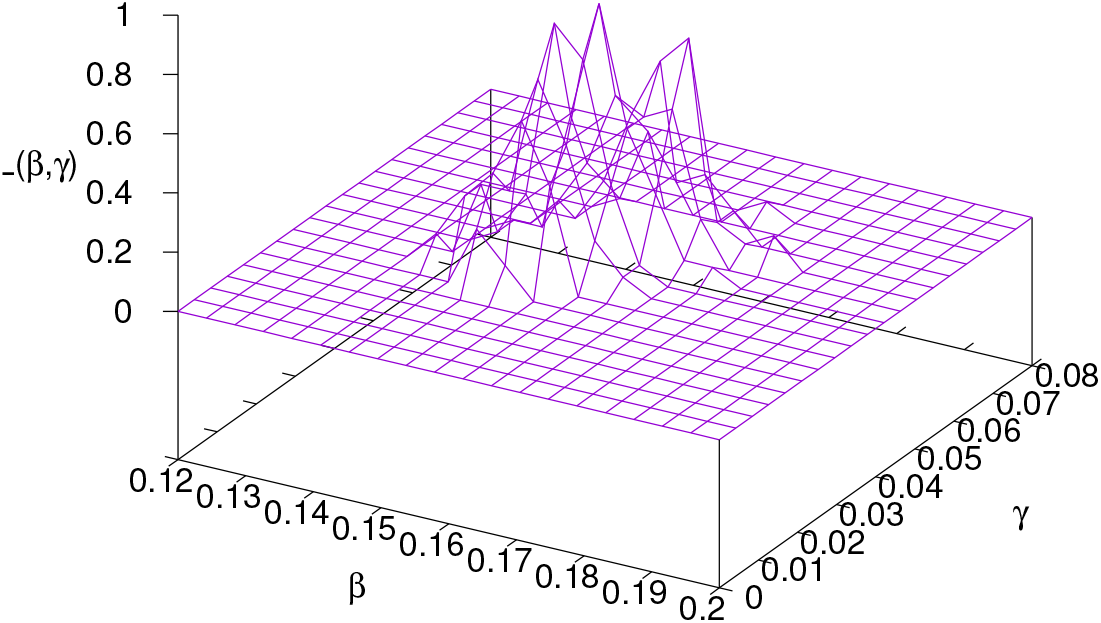
Two-parameter likelihood plot *L*(*β, γ*) for parameters the infection rate *β* and the recovery rate *γ*, showing fewer than expected correlations between these two parameters which essentially determin the common growth factor *λ*_1_. This well determined range of possible values for the parameters *β* and *γ* might be due to the available information from all 5 data sets, including the recovered in good quality.

Furthermore, combinations between the parameters describing the distiction between severe and mild infecteds could be expected to show large insecurities in the individual parameters as well as large correlations between combinations of them. Hence we investigate in numerical two parameter plots the combinations of the severity ratio *η* with the infectivity ratio of mild infected as compared to the severely diseased, the ratio *ϕ*, hence *L*(*η, ϕ*), and the detection ratio *ξ* of mild or asymptomatic, hence *L*(*η, ξ*), and further the likelihood of the two last mentioned parameters *L*(*ϕ, ξ*), see Fig. 12.

**Figure 12:**
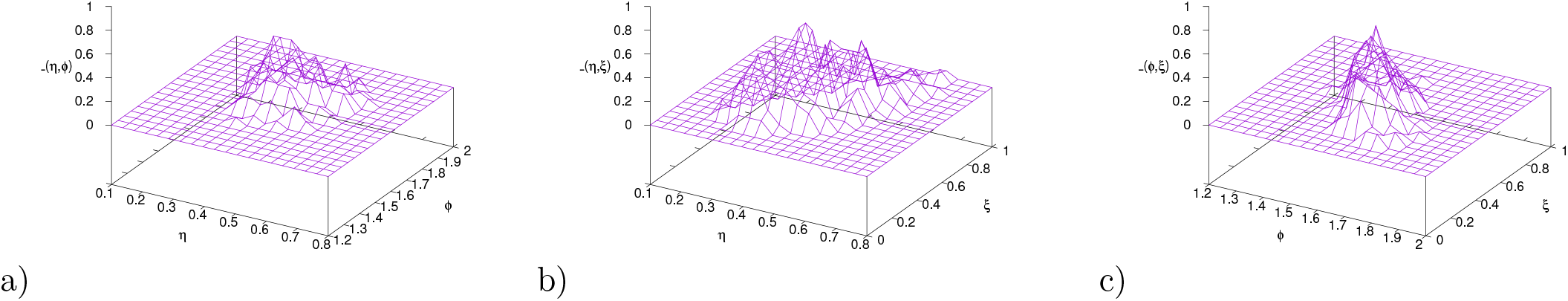
Two-parameter likelihood plot a) for *L*(*η, ϕ*), b) for *L*(*η, ξ*), and c) for *L*(*ϕ, ξ*), the parameters which describe more internal effects of hospitalization ratio *η* versus asymptomatic/mild, and infectivity of mild/asymptomatic *ϕ*, and detection rate of mild/asymptmatic *ξ*. Largest correlations between parameters are observed between *η* and *ξ*, see Fig. part b).

The likelihood *L*(*η, ϕ*), Fig. 12 a), is well in a range close to what we would expect form the individual likelihood plots in Fig. 3 e) and f). The largest correlations are visible in the plot for *L*(*η, ξ*), Fig. 12 b), where we observe non-vanishing probabilities of parameters for small hospitalization rate *η* and small detection rate of mild/asymptomatic infections *ξ* as well as for large *η* and large *ξ*. However, the severity rate *η* is well restricted between 0.2 and 0.7 and a clear maximum visible around the best value from the individual liklihood plot, Fig. 3 e). The boundary value of *ξ* = 1 has a non-vanishing probability to explain the data, but numerically the value of *L* there is much smaller than the values in the middle of the parameter intervals of *η* and *ξ*. Hence, even with higher numerical resolution, on the expense of many more stochastic runs of the model, it is not expected to have high values of *L*(*η, ξ*) still at locations close to the bundaries of the possible parameter values. So there are considerable correlations between these two parametes detected, but they seem to be mild, and from the available data we can obtain good information also about the most likely values of these parameters. Finally, for the parameter combination of *L*(*ϕ, ξ*) we observe again a more restricted area of possible parameter sets, though also here some correlations are visible. In conclusion, it seems that the available data sets give quite some information about the possible parameter combinations to describe the system under investigation. Further refinements of these analyses might give further insight into the dynamics of the epidemic.

Due to initially small numbers of infected, hospitalized cases etc., we use initially the Gillespie algorithm for the state discrete Markov process described by the master equation [37, 29]. The present two parameter likelihood plots become occasionally very computationally demanding, depending on system size *N* and numbers of transitions, especially for larger infectivity. hence the well tested approximation via the Fokker-Planck equation as stochastic differential equation system [40] speeds up the simulations, allowing for smoother likelihoods, but occasionally might give some errors in the small number situations. A carefull monitoring is therefor needed, see a first comparison of master equation likelihoods, using Eq. (3), with stochastic differential equation likelihoods, using Eq. (16), with encouraging results Fig. 13, but need to be further investigated in future studies. However, the first results given here in Figs. 11 and 12 are already informative for the purpose of detecting eventual correlations between parameters, and encouraging for further intensive studies. The up to now detected correlations keep well within the expected ranges indicated from the one parameter numerical likelihood plots in Fig. 3.

**Figure 13:**
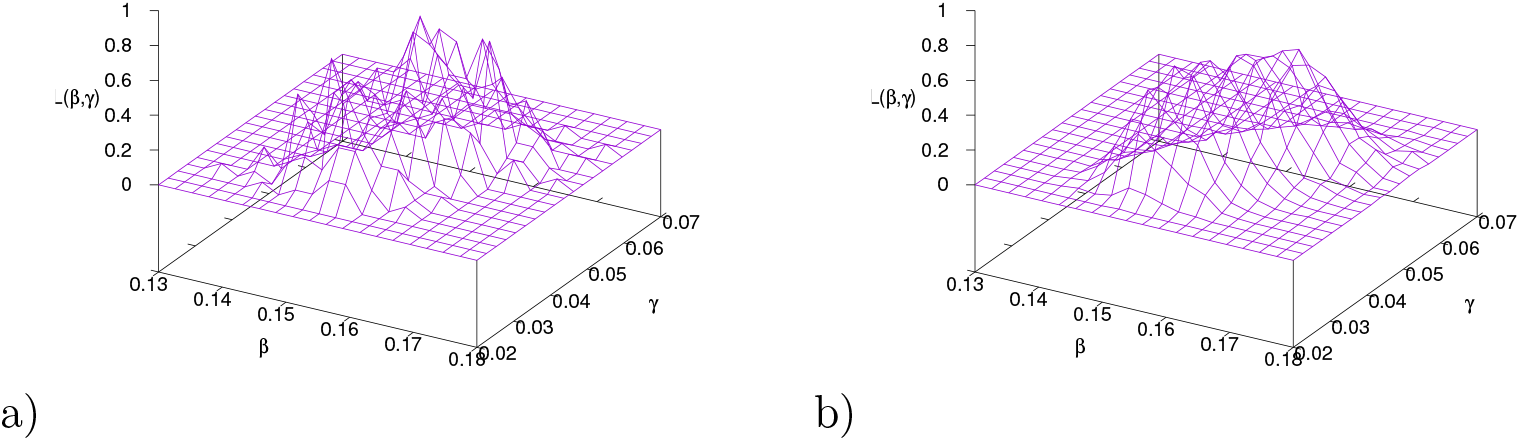
Two-parameter likelihood plots for *L*(*β, γ*) zooming into the parameter regions of *β* and *γ*, in a) comparing ensembles from the master equation simulations via Gillespie algorithm with the empirical data, and in b) using ensembles from the Fokker-Planck approximation simulating the stochastic differential equation system. The elevated areas of both graphs show a similar parameter region, confirming that the original approach a) already gives good informations about the two-parameter likelihood, and refinements in b) given a smooth likelihood surface, since larger ensembles are now better accessible.

